# Large-scale functional characterization of low-density lipoprotein receptor gene variants improves risk assessment in cardiovascular disease

**DOI:** 10.1101/2023.12.27.23299827

**Authors:** Mohammad Majharul Islam, Max Tamlander, Iryna Hlushchenko, Samuli Ripatti, Simon G. Pfisterer

**Affiliations:** Department of Anatomy, Faculty of Medicine, University of Helsinki, Finland; Institute for Molecular Medicine Finland, FIMM, HiLIFE, University of Helsinki, Helsinki, Finland; Clinicum, Department of Public Health, University of Helsinki, Helsinki, Finland; Broad Institute of MIT and Harvard, Cambridge, MA, USA

**Keywords:** Low-density lipoprotein receptor (LDLR), familial hypercholesterolemia (FH), whole exome sequencing, rare variant, cardiovascular disease, hypercholesterolemia

## Abstract

**Aims:** Lack of functional information for low-density lipoprotein (LDL) receptor (LDLR) mutations limits the use of genetics for early diagnosis, risk assessment and clinical decision making in familial hypercholesterolemia (FH). The goal of this study was an in-depth and large-scale functional characterization of *LDLR* variants to overcome this problem.

**Methods:** Open-source robotic tools were integrated with multiplexed high-content microscopy, image and data analysis into a novel semi-automated analysis pipeline for the characterization of *LDLR* variants to quantify LDL uptake, LDLR localization and expression.

**Results:** 315 *LDLR* coding variants were functionally characterized in this study and collapsed into four functional groups based on their residual LDL uptake activity (“Loss-of-function”, 0-10% activity; “defective”, 10-30%; “mildly-defective”, 30-70%; and “non-defective”, > 90%). Integration of the activity groups with whole-exome sequencing and clinical data from UK biobank demonstrated that considering LDLR activity levels improved risk assessment in dyslipidaemia and cardiovascular disease (CVD). Individuals carrying *LDLR* variants from the loss-of-function and defective groups displayed increased odds ratios for CVD (OR=6.1, 95% CI = 1.5 - 24.4; OR = 1.83, 95% CI = 1.2 - 2.7) as compared to the non-defective group. Also, plasma LDL-cholesterol, utilization of lipid-lowering drugs and combination therapy were higher in the loss-of-function (OR = 15.4, 95% CI = 3.8 - 61.7; OR = 7.6, 95% CI = 1.8 - 31.8; OR = 96.8, 95% CI = 22.6 - 414.1), defective (OR = 5.9, 95% CI = 4.1 - 8.6; OR = 3.5, 95% CI = 2.5 - 4.9; OR = 15.6, 95% CI = 8.4 - 29.1) and mildly-defective group (OR = 2.0, 95% CI = 1.5 - 2.7; OR = 2.0, 95% CI = 1.6 - 2.4; OR = 1.9, 95% CI = 1.0 - 3.4) as compared to the non-defective group. Especially, the loss-of-function group displayed higher CVD risk, increased LDL-C and combination therapy usage as compared to the ClinVar pathogenic group for the same subjects. Furthermore, the functional data indicates that prediction tools tend to overestimate the fraction of pathogenic *LDLR* variants.

**Conclusion:** Systematic functional data for *LDLR* variants paves the way for improved diagnosis, risk assessment and treatment optimization for FH patients, enabling a better utilization of genetic data in clinical decision making.

**Translational perspective:** A loss-of-function *LDLR* variant leads to lifelong exposure of elevated LDL-C. Whilst sequencing of the *LDLR* gene is included in the genetic assessment of FH patients, most *LDLR* variants lack information about functional consequences at the cellular level. This limits the utility of genetic tools in the diagnosis and treatment of FH. This study overcomes this problem, providing functional information for a large set of *LDLR* variants. Integration with genetic and clinical data from UK biobank enables links between functional and clinical effects, making it easier to diagnose FH and estimate a patient’s cardiovascular risk.

## Introduction

Elevated low-density lipoprotein cholesterol (LDL-C), hypercholesterolaemia, leads to cardiovascular disease, one of the most common causes of death worldwide^1,2^. Familial hypercholesterolemia (FH) manifests a severe form of hypercholesterolaemia, with up to 20 fold higher risk for cardiovascular disease (CVD) and severely increased LDL-C levels as compared to healthy individuals^3,4^. FH is considered a monogenic autosomal dominant disorder with mutations in *LDLR*, *APO-B* and *PCSK9*^5,6^.

Only less than 10% of FH patients are diagnosed^7^. Moreover, most FH patients do not achieve their LDL-C target levels^3,7^. Genetic testing offers great promise for detecting and treating FH patients at an early stage. However, the applicability of genetic testing is hampered by a lack of functional data. More than 85% of *LDLR* variants are functionally uncharacterized^8^ and the speed of functional testing is slow with the largest studies reporting functional activities for 46 and 68 variants^9,10^. Assessing the pathogenicity of a variant mostly relies on correlations with a clinical phenotype, allele frequency and prediction tools to estimate variant effects^6^.

Especially rare variants are difficult to assess with this approach. This limits the applicability of genetic testing for early diagnosis of FH and complicates the interpretation of *LDLR* variant effects. Utilization of functional activity scores for *LDLR* variants, might provide additional insight for precision medicine applications as compared to a binary grouping into pathogenic or benign variant groups.

Deciphering the impact of genetic mutations on cellular processes is time consuming. Several methods have been brought forward to solve this problem, including saturation genome editing^11,12^, deep-mutational analysis^13^, or base-editor screens^14,15^. However, these methods rely on readouts which can be traced easily, such as cell viability or large differences in fluorescence signal intensities. So far, we lack technologies that enable multiplexed cellular readouts, delivering multifaceted insight into functional defects of a variant and providing an activity range for the functional defects of each variant as compared to the wild type.

To overcome current challenges in the systematic functional characterization of genetic variants, we set out to integrate open and affordable robotic systems with automated multiplexed high-content microscopy and CRISPR-based gene editing. This resulted in a semi-automated and scalable workflow which yields multiplexed functional data for individual genetic variants. As a proof-of-concept, we applied this strategy to *LDLR* coding variants, characterizing 315 variants. We linked the functional cell data with whole-exome sequence sequencing data from the UK biobank and demonstrated the added benefit of functional variant characterization and the resulting activity groups for risk assessment and clinical decision making in FH.

## Methods

Reagents, cell culture procedures, generation of LDLR knockout cells, establishment of *LDLR* variant cell lines, fluorescent LDL uptake experiments and image analysis are explained in the Supplemental Methods section.

### Data processing and statistical analysis

Data processing and visualization were conducted using the standard Python libraries (www.python.org) and the following packages: pandas, numpy, scipy, matplotlib, and seaborn. The single-cell level raw data of *LDLR* variants were initially normalized against *LDLR WT* for each experiment and condition. The normalized data was then combined to generate treatment-level mean DiI-LDL intensity, GFP intensity, and DiI-LDL positive organelle counts per cell. The mean DiI-LDL intensity (DiI-Mean) and DiI-LDL positive organelle counts (DiI-Orgs) were then averaged to determine the residual activity of each variant. Residual activity of variants located in the ligand-binding domain was further corrected using their expression levels. Statistical analyses were performed with Python packages. To compare the pairwise effect for variant groups, statistical significance was determined using the Mann-Whitney U test. Fisher Exact tests were used to calculate the statistical significance for odds ratios.

### UK Biobank cohort

Clinical data of UK Biobank participants carrying the *LDLR* coding sequence variants were retrieved utilizing their whole-exome sequence (WES) data (Final release). Of the 315 variants characterized in the current study, 124 were identified in the UK Biobank cohort, comprising a total of 11,797 participants carriers. Individuals with records for statin usage were defined as being on lipid-lowering medication. Participants taking additional lipid-lowering drugs such as: Ezetimibe, bile acid sequestrants or fibrates were defined as being on combination therapy. CVD were defined as either myocardial infarction, stroke, angina and ischemic attack and collected using UK Biobank field codes: 6150 (Vascular/heart problems diagnosed by doctor), 41271 (Diagnoses - ICD9), 41270 (Diagnoses - ICD10) and 20002 (Non-cancer illness code, self-reported). Information provided during the enrollment period was considered for analysis.

### Fluorescent LDL uptake assay

Cells were seeded into 384-well plates using an OT-1 (Opentrons) robotic platform. Cells were seeded in triplicates for each treatment condition. Initial seeding was performed in lipid-rich (LR) medium supplemented with 10% FBS. After 24 hours, the medium was replaced with lipid-poor (LP) medium containing 5% LPDS for wells designated for LP and LS treatments. After 48 hours, wells designated for lipid-poor plus mevastatin (LS) treatment received LS medium containing mevastatin at a concentration of 10 μg/ml. Following 72 hours of overall treatment, DiI-LDL uptake was stimulated by adding 10 μg/ml DiI-LDL solution and incubated for 30 minutes at 37°C. Subsequently, cells were rinsed with phosphate-buffered saline (PBS) and fixed with 4% paraformaldehyde (PFA). Subsequently, cells were washed and stained with Hoechst 33342 (5 μg/ml) and CellMask Deep Red (0.5 μg/ml). Images were acquired using a PerkinElmer OperaPhenix automated spinning disc confocal microscope equipped with a 20x or 40x water immersion objective. For each well, five image stacks with four layers were acquired. Each LDL uptake experiment contained four control cell lines: wild-type HepG2, LDLR KO, and two LDLR WT-GFP expressing KO cell lines. Two independent LDL uptake assays were performed for each LDLR variant cell line.

## Results

### Human cell system to study *LDLR* gene variants in a scalable fashion

We established a semi-automated platform for reliable and systematic quantification of *LDLR* variants. For this purpose, we selected the HepG2 cell line, which is derived from liver origin and is amenable for genetic modification with CRISPR technology.

First, we produced a HepG2 LDLR knockout cell line (LDLR KO) with CRISPR, derived from a single cell clone (Fig. 1a), and verified the cell line by DNA sequencing (Supplementary Fig. 1a) and high-content microscopy of surface and cellular (surface + internal) LDLR expression (Supplementary Fig. 1b,c). LDL uptake studies were performed as additional validation of LDLR deficiency. Fluorescent LDL particles (DiI-LDL) were added to the cells after preincubation with lipid-rich medium (LR) (10% fetal bovine serum, (FBS), lipid-poor medium (LP) (5% lipoprotein depleted serum, (LP)) and lipid-poor medium including mevastatin (LS) to block cholesterol synthesis (Fig. 1b, c). Mean cellular DiI-LDL intensities (DiI-Mean) and the number of DiI-LDL positive organelles per cell (DiI-Org) were quantified by automated high-content imaging. Whilst the quantification of DiI-Org reflects internalized LDL, DiI-Mean combines signals from internalized and surface bound LDL particles. In wild-type cells DiI-Mean intensities (LR 0.13, LP 0.39 and LS 0.91) and DiI-Org numbers (LR 3.5, LP 16.4, and LS 31.2) increased in a stepwise manner from lipid-rich to lipid-poor plus mevastatin conditions (Fig. 1c). However, in LDLR KO cells LDL uptake was blunted as visualized by DiI-Mean (LR 0.009, LP 0.007, LS 0.013) and DiI-Org (LR 0.09, LP 0.12 and LS 0.14) quantifications.

**Figure 1:**
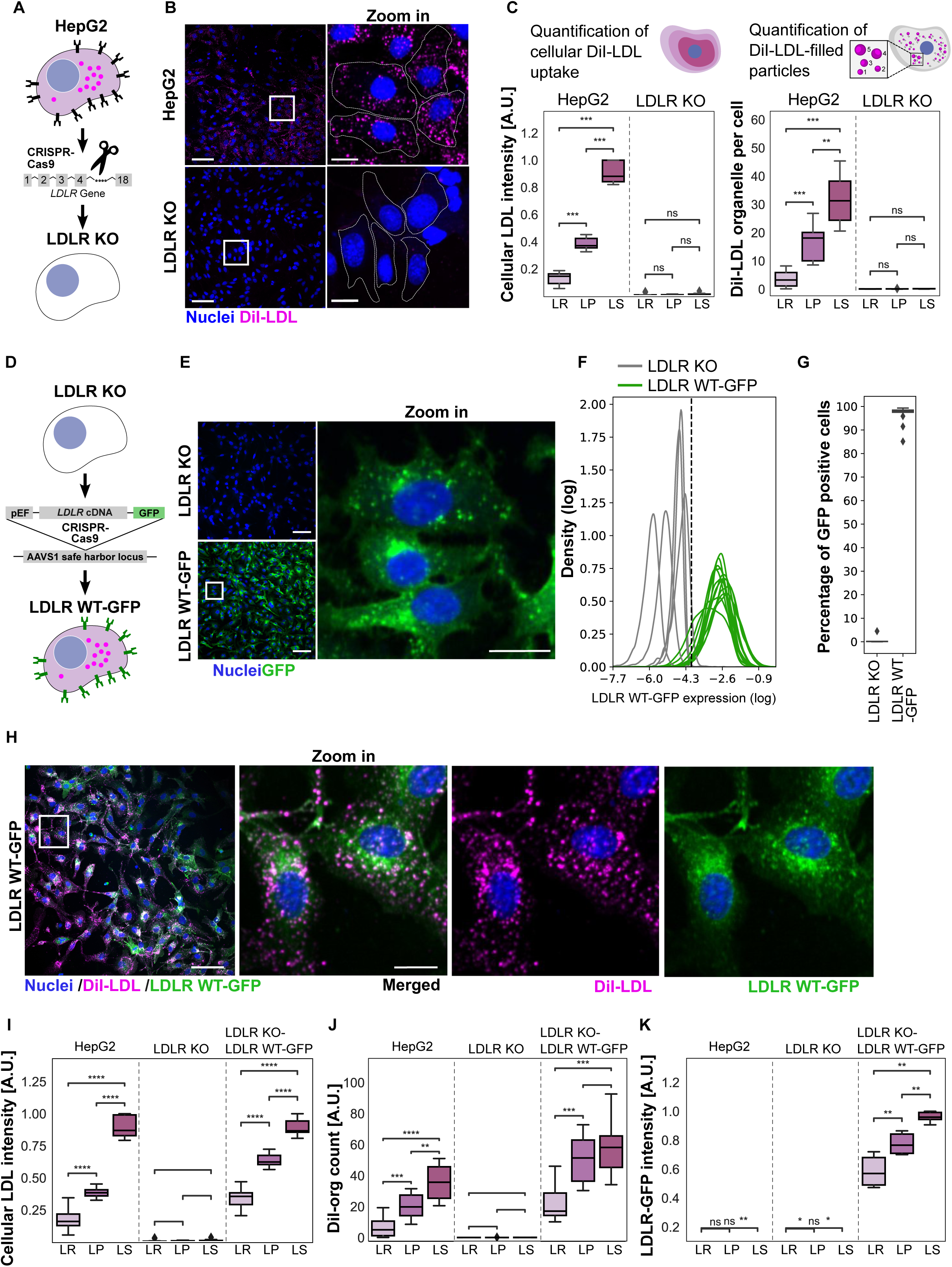
HepG2 cell model for characterization of LDLR variants. (A) Schematic presentation of generating a LDLR knockout cell line using CRISPR-Cas9. (B) Representative image fields showing DiI-LDL uptake in HepG2 wild-type (WT) and LDLR knockout (KO) cells in lipid poor conditions. (C) Quantification of mean DiI-LDL intensity and average number of DiI-LDL filled organelles per cell in HepG2 WT and LDLR KO cells in three treatment conditions, lipid rich (LR), lipid poor (LP) and lipid poor plus mevastatin (inhibition of cholesterol synthesis)(LS). Quantitative analysis was carried out on over 10000 cells from three independent experiments. (D) Schematic illustration depicting the stable reintroduction of the LDLR-WT-EGFP gene into the genome of LDLR KO cells at the AAVS1 locus. (E) Representative image field showing stable expression of LDLR WT-GFP in LDLR KO cells. (F) Logarithmic density plot displays EGFP signal intensities for KO-LDLR-WT-GFP expressing and LDLR KO cell lines. The dashed line indicates the threshold value for discriminating GFP-positive from GFP-negative cells. (G) Box plot for GFP-positive and GFP-negative cells quantified from LDLR KO and LDLR WT-GFP expressing cell lines, with quantification of over 15000 cells from five independent experiments. (H) Representative image field demonstrating the restoration of DiI-LDL uptake in LDLR-WT-GFP expressing knockout cells with zoom in of the indicated area for DiI-LDL and LDLR-GFP channels. Quantification of mean DiI-LDL intensity (I), average DiI-LDL filled organelles per cell (J) and mean LDLR WT-GFP intensity (K) in HepG2 WT, LDLR KO and LDLR WT-GFP cells in lipid-rich, -poor and lipid poor plus statin conditions. Over 15000 cells from three independent experiments were quantified. Statistical significance was determined using the Mann-Whitney U test, with asterisks denoting significance levels: *p < 0.05, **p < 0.01, ***p < 0.001 and ****p < 0.0001.

To achieve uniform expression of LDLR variants in a large number of LDLR KO cells we went forward with a stable expression system reported previously^16^, which allowed us to integrate LDLR-GFP expression cassettes into the genome at the AAVS1 safe harbor locus with CRISPR/Cas9 (Fig. 1d). Stable LDLR-GFP expressing cells were enriched with puromycin selection. Inspection of high-content images verified a uniform LDLR-GFP expression pattern (Fig. 1e), with LDLR-GFP detectable at the plasma membrane and in intracellular organelles, reflecting previously reported localizations of LDLR (Fig. 1e)^17^. Plotting LDLR-GFP intensities for individual cells from several independent LDLR-GFP cell lines further verified that LDLR-GFP was expressed at comparable levels and was distinct from non-expressing LDLR KO cells (Fig. 1f). We selected the 3rd percentile of the LDLR-GFP intensity as a cut-off value to distinguish LDLR-GFP positive from negative cells (Fig. 1f). Across five newly generated LDLR-GFP wild-type cell lines, 97% of the cells were positive for LDLR-GFP (Fig. 1g).

LDLR-GFP colocalized with DiI-LDL in cytoplasmic foci resembling endosomal organelles (Fig. 1h) and reestablished LDL uptake activity in LDLR KO cells (Fig. 1h,i,j). In lipid-rich conditions DiI-Mean intensities and DiI-Orgs were higher in LDLR KO cells expressing LDLR-GFP wild-type (KO-LDLR-WT-GFP) as compared to HepG2 (0.33 and 0.17 for DiI-Mean; 22.3 and 7.2 for DiI-Orgs), likely due to low level constitutive expression of LDLR-GFP. Mean-DiI and DiI-Org further increased in lipid poor (DiI-Mean 0.66, DiI-Orgs 51) and lipid poor plus mevastatin (DiI-Mean 0.96, DiI-Orgs 58) in KO-LDLR-WT-GFP cells (Fig. 1i,j). This was paralleled with a concomitant increase in cellular LDLR-GFP intensity in lipid poor and lipid poor plus mevastatin conditions as compared to lipid rich conditions (LR 0.57, LP 0.79 and LS 0.97 (Fig. 1k).

### Large-scale generation of *LDLR* variants and introduction into LDLR KO cells

So far, functional assays for *LDLR* variant characterization were limited in throughput. We set up a semi-automated pipeline for the generation of expression constructs and introduction into LDLR KO cells. In a first step, expression constructs were made with Gibson assembly^18^: For each variant two LDLR fragments containing the desired mutation were amplified with PCR, purified and assembled with the vector backbone (Fig. 2a). Then, PCR reactions, fragment isolation, concentration adjustment, and plasmid DNA isolations were automatically pipetted with an open-source robotic platform (Fig. 2a). More than 350 expression constructs were made, of which 98% contained a correctly assembled LDLR-GFP cDNA (Fig. 2a). Then we validated the introduction of the desired mutation with a customized next-generation sequencing approach for 214 variants (Fig. 2b). 150 bp stretches surrounding the desired mutation were PCR amplified, barcoded, pooled and submitted to next-generation sequencing. This allowed us to confirm 91.5% of *LDLR* variant constructs, 6% lacked the desired mutation and 1.3% displayed either an alternative or an additional mutation. These *LDLR* variants were removed from the analysis (Fig. 2b).

**Figure 2:**
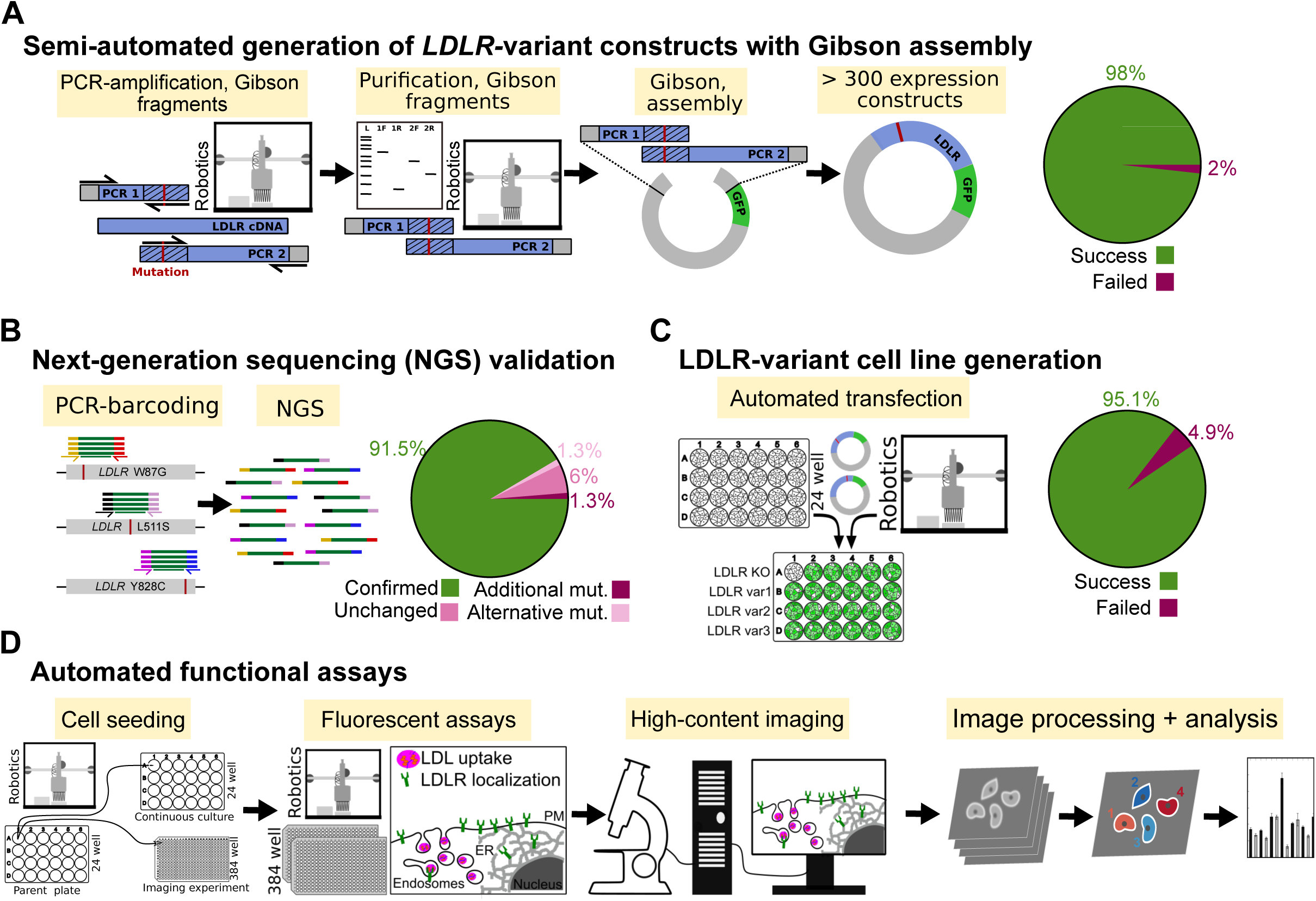
Semi-automated pipeline for the construction of expression constructs and characterization of LDLR variants. (A) Schematic presentation of generating LDLR variant constructs. (B) Schematic illustration of the next-generation sequencing workflow for LDLR variant verification, with a pie chart summarizing the success rate for LDLR variant generation. (C) Depiction of automatic transfection and its efficiency in generating LDLR variant cell lines. (D) A schematic presentation of semi-automated cell seeding in 384 well plates, treatment with different lipid starvation conditions, DiI-LDL labeling, cell fixation and staining, high-content fluorescence microscopy, image processing, and image and data analysis for evaluating functional consequences of LDLR variants.

Semi-automated cell culture techniques were established on an open-source robotic platform to transfect LDLR KO cells with *LDLR* variant and CRISPR expression constructs (Fig. 2c). Stable *LDLR* variant expressing cell lines were obtained successfully for 95% of the *LDLR* variant constructs (Fig. 2c). Mean-GFP intensities for individual *LDLR* variants were distinct from non-expressing LDLR KO cells, but more variable as compared to wild-type LDLR-GFP (Supplementary Fig. 1d). Open-source robotics were used to perform LDL uptake experiments and to prepare cells for high-content imaging in semi-automated fashion. Images were automatically acquired with a spinning-disk confocal microscope, quantified with CellProfiler and analyzed with Python (Fig. 2d). With this approach we characterized 315 *LDLR* variant cell lines in two independent experiments, in lipid rich and lipid poor conditions. This reflects data from more than 5400 wells of a 384 well plate and 2.2 million cells. Data for individual *LDLR* variants is contained in Supplementary Table 1.

### Functional characteristics of 315 *LDLR* variants

On average, functional profiling was performed with 12 *LDLR* variant cell lines per experiment together with HepG2 wild-type, LDLR KO and KO-LDLR-WT-GFP cell lines as controls. DiI-Mean, DiI-Org and Mean-GFP values from a variant cell line were normalized to the respective readouts from the KO-LDLR-WT-GFP cell line. This allowed us to obtain an activity score for each variant and to compare *LDLR* variant quantifications from different experiments. As an overview, we displayed normalized DiI-Mean, DiI-Org and Mean-GFP values for each *LDLR* variant in lipid-rich and poor conditions, grouped by their position in LDLR domains (Fig. 3a). *LDLR* variants with reduced activity were observed in all domains. Interestingly, *LDLR* variants from the ligand-binding domain exhibited higher expression levels compared to variants from other domains (Fig. 3a, Supplementary Fig. 2a)

**Figure 3:**
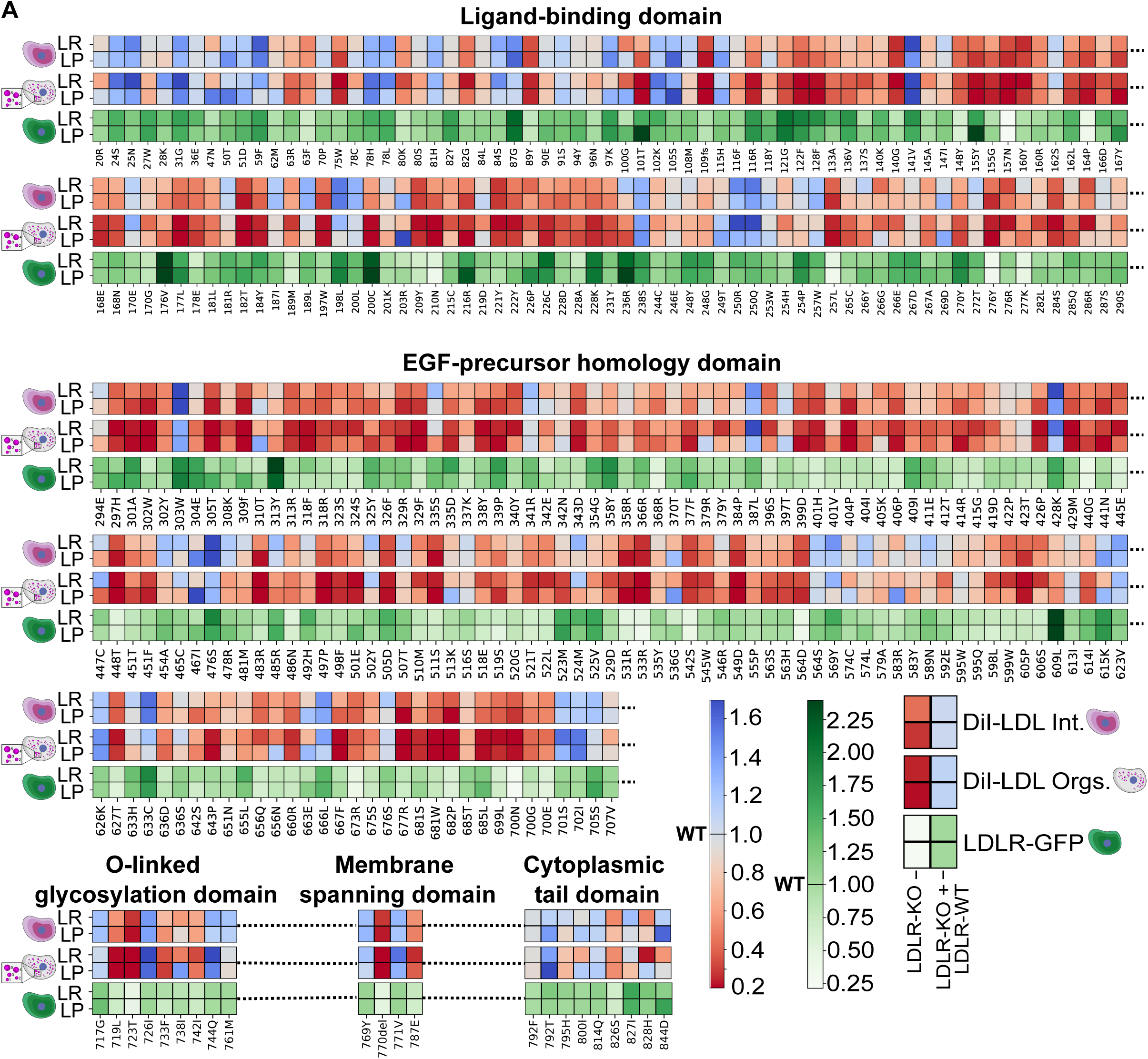
Heatmap depicting the functional activity and expression of 315 LDLR variants relative to wild-type LDLR in lipid-rich and lipid-poor conditions. The presented values for individual LDLR variants were normalized to LDLR-WT-GFP in regards to mean DiI-LDL intensity, DiI-LDL organelle counts, and expression levels of the variant constructs. The variant information was sorted according to their position in the LDLR domains.

To categorize *LDLR* variants into functional activity groups, we combined DiI-Mean and DiI-Org readouts in lipid poor conditions from two independent experiments. In addition, the DiI-Mean and DiI-Org readouts of variants from the ligand-binding domain were normalized for LDLR expression. According to the Clinical Genome Resource (ClinGen, www.clinicalgenome.org) FH variant curation expert panel (VCEP) guidelines^6^, variants with less than 70% residual activity should be grouped as pathogenic, whilst those with an activity above 90% would correspond to benign variants. Considering this classification, 63% (198) of the variants were pathogenic and 22% (72) benign (Fig. 4b). We splitted the group of *LDLR* variants with less than 70% activity into additional groups to assess whether different levels of residual LDLR activity have an impact on disease progression (Fig. 4c). *LDLR* variants with 0-10% residual activity were labeled as “loss-of-function”, those with 10-30% as “defective”, and variants with 30-70% as “mildly-defective” (Fig. 4c).

**Figure 4:**
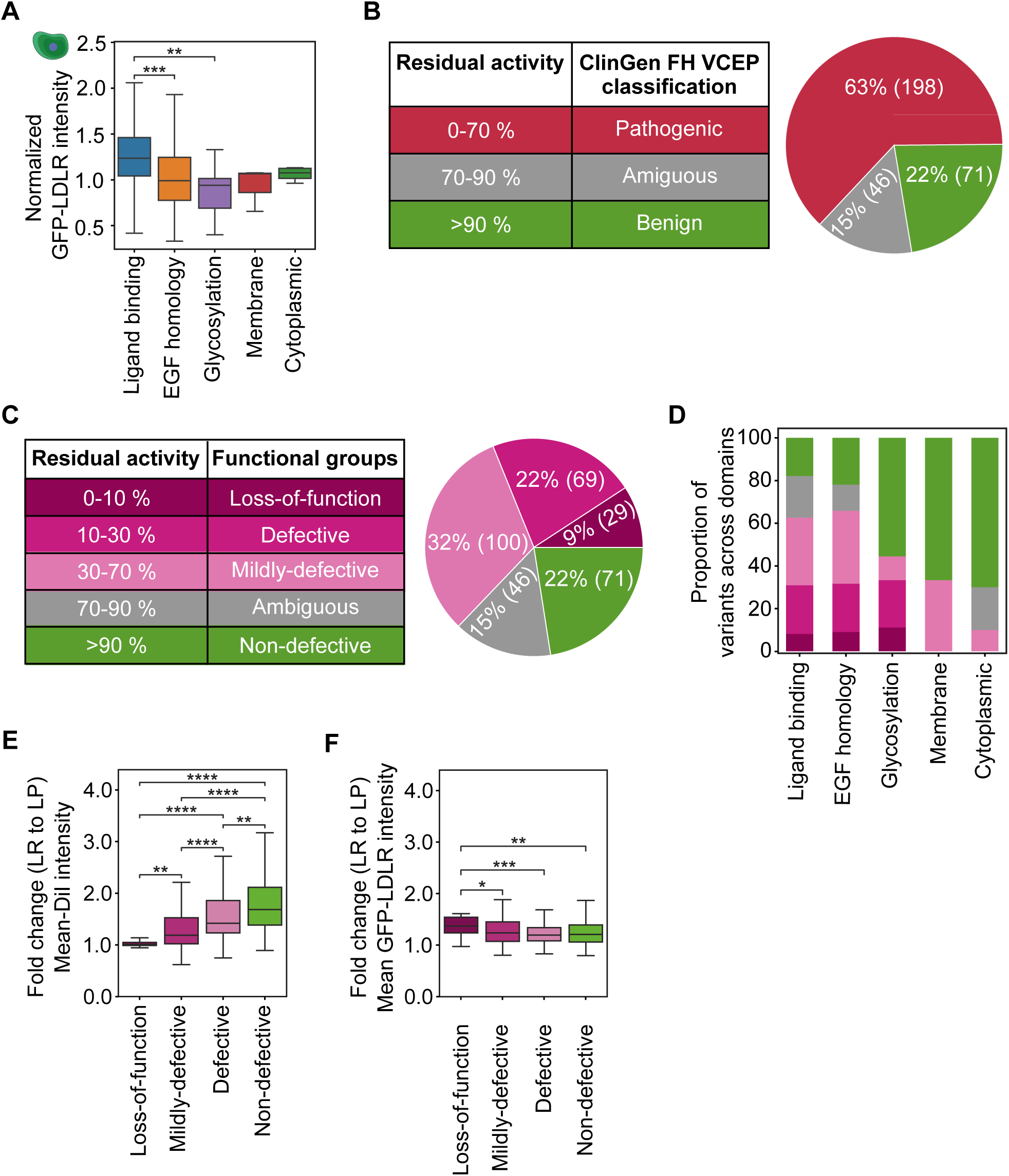
Categorization of LDLR variants based on their residual LDL uptake activity computed from mean cellular DiI-LDL intensity, DiI-LDL organelle counts and GFP expression of the respective variant. (A) Quantification of LDLR variant GFP expression across LDLR domains. (B) Classification of LDLR variants based on their residual LDL uptake activity. (C) Relative distribution of LDLR activity groups among different LDLR domains. (D) Quantification of the increase in mean cellular DiI-LDL intensity for LDLR variants in response to the transition from lipid-rich to lipid-poor conditions expressed as fold change by dividing the results from lipid-poor conditions by the results obtained from lipid-rich conditions. Grouped based on the residual LDL uptake activity. (E) Quantitative determination of the fold change in mean cellular GFP expression for LDLR variants in response to the transition from lipid-rich to lipid-poor conditions grouped by the residual LDL uptake activity. Statistical significance was determined using the Mann-Whitney U test, with asterisks denoting significance levels: *p < 0.05, **p < 0.01, ***p < 0.001 and ****p < 0.0001.

Of the 315 LDLR variants, 9% represented loss-of-function, 22% defective, 32% mildly-defective, 15% ambiguous and 22% non-defective groups (Fig. 4c). The fraction of loss-of-function and defective variants was highest in the ligand-binding domain, the EGF-homology-precursor and the o-linked glycosylation domain (Fig. 4d) as compared to the membrane spanning and cytoplasmic tail domains (Fig. 4d). The ability to upregulate LDL internalization in lipid-poor conditions was decreased for variants of the loss-of-function group and increased in a stepwise manner for variants of defective, mildly-defective and non-defective groups, as quantified by the fold increase in DiI-Mean intensity in lipid-poor versus rich conditions (Fig. 4e). On the other hand, increased LDLR expression in lipid-poor conditions was more profound in the loss-of-function group as compared to other LDLR activity groups (Fig. 4f).

### Comparing functional data with computational tools and ClinVar classification

To predict the functional consequences of *LDLR* variants, we utilized some of the most commonly used tools: Sift^19^, MutationTaster^20^, PolyPhen2^21^ and PHD-SNPg^22^. A total of 148 *LDLR* variants included in our study were labeled pathogenic by all of the prediction tools employed (Fig. 5a). However, this seems to be an overestimation as only 15.5% (23) of the predicted pathogenic variants were quantified as loss-of-function and 28.4% (42) as defective using our functional approach (Fig. 5b). The predicted pathogenic variants contained 9.5% (14) of non-defective, 9% (13) ambiguous and 37.8% (56) mildly-defective variants (Fig. 5b). Next, we visualized how our functional data compared to existing ClinVar classification of the same *LDLR* variants using a flow diagram (Fig. 5c). Variants listed as pathogenic in ClinVar did not contain any “non-defective” variants and showed the highest percentage of loss-of-function variants (33%) (Fig. 5c,d), which decreased to 12% in the likely-pathogenic group (Fig. 5d).

**Figure 5:**
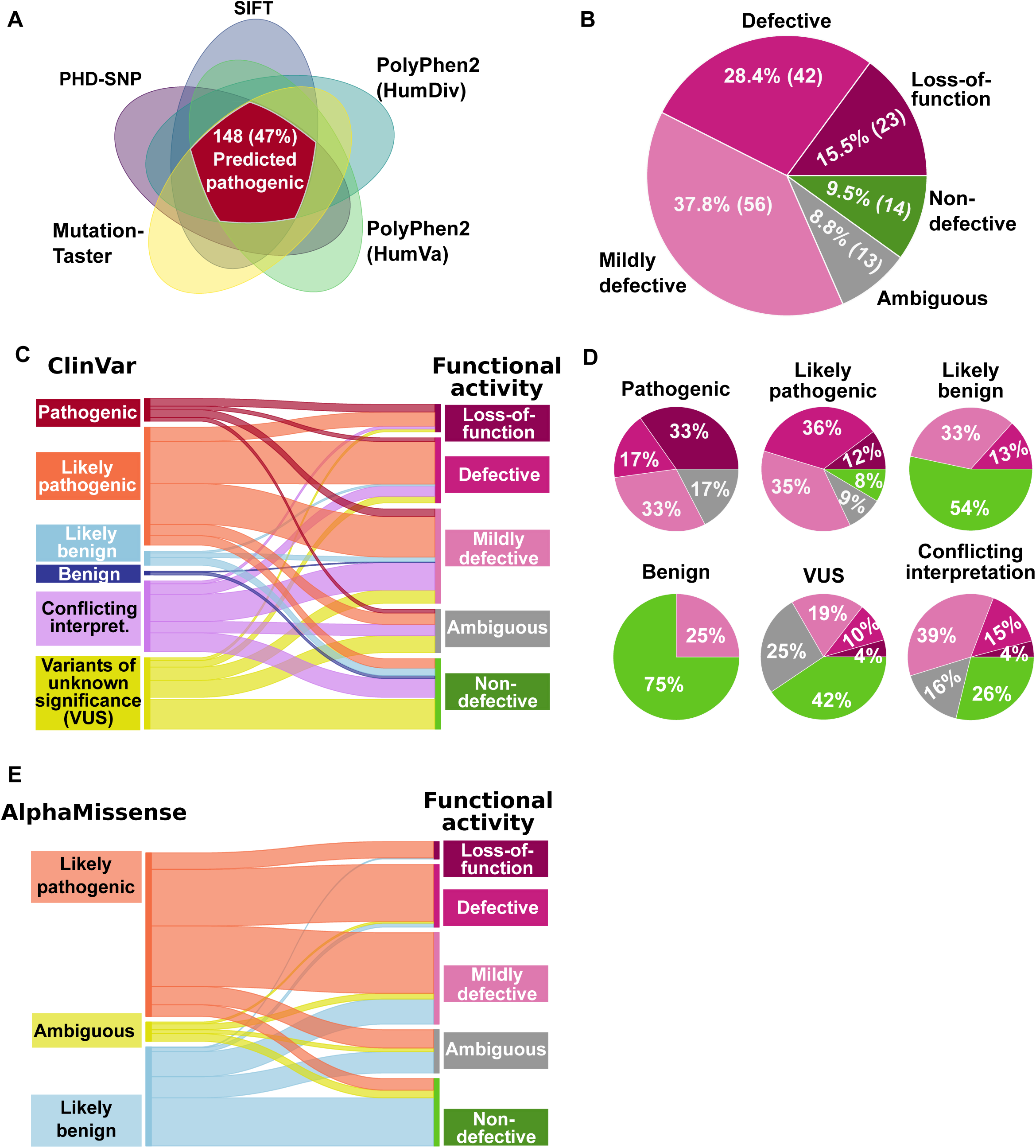
Comparison of functional activity groups and established LDLR variant classifications. (A) Graphical illustration of predicted pathogenic LDLR variants identified by five different prediction software tools. (B) Pie chart demonstrating the segregation of predicted pathogenic LDLR variants into distinct functional activity groups. (C) Flow diagram illustrating the segregation of LDLR variants from functional activity groups as compared to ClinVar classification. (D) The pie charts illustrate the distribution of LDLR variants from different activity groups within individual ClinVar classes. (E) A flow diagram depicts the channeling of LDLR variants from different AlphaMissense classes towards distinct functional classes. (F) Assigning ClinGen classification, a set of standardized guidelines established by an expert panel for Familial hypercholesterolemia variants, based on their functional scores.

Loss-of-function variants were undetectable in the likely-benign and benign groups (Fig. 5d). Defective *LDLR* variants were contained in the pathogenic (17%), likely-pathogenic (36%) and likely-benign (13%) groups but not in the benign group. Vice-versa, non-defective variants first appeared in the likely-pathogenic group (8%) and then increased in the likely-benign (54%) and benign (75%) groups (Fig. 5d). We also compared our functional data with results from AlphaMissense which sorts variants into likely pathogenic, likely benign and ambiguous groups^23^. Also in this case predicted likely-pathogenic variants contained a fraction of non-defective, mildly-defective, and ambiguous *LDLR* variants. Except for one, all loss-of-function variants were contained within the likely-pathogenic group (Fig. 5e).

### Integration of whole-exome sequencing and clinical data from UK Biobank with functional *LDLR* variant groups

For evaluation of our classification with clinical phenotypes we used our functional *LDLR* variant groups and integrated them with whole-exome sequencing data, clinical, biochemical and NMR metabolomics data for 502 364 individuals from the UK biobank. 124 of the functionally analyzed *LDLR* variants were contained in UK Biobank. 5 variants and 8 subjects were identified for the loss-of-function group, 15 variants and 117 subjects for the defective group, 39 variants and 1165 subjects for the mildly-defective (30-70%) group and 65 variants and 10487 subjects for the non-defective group (Fig. 6a). Loss-of-function carriers displayed an OR of 15.4 (CI: 3.8-61.7, p < 0.001) to have a circulating LDL-C above 5 mmol/l as compared to the non-defective variant group. Also the OR for the defective group (5.9, CI: 4.1 - 8.6, p < 0.001) and carriers of mildly-defective variants (1.6, CI: 1.3-2.0 p < 0.001) were higher (Table 1). The effects of the different functional activity groups on circulating LDL-C were also visible in box plots (Supplementary Fig 2b-h), with the loss-of-function group resulting in higher plasma LDL-C, total cholesterol and APO-B than the defective and mildly defective groups.

**Figure 6:**
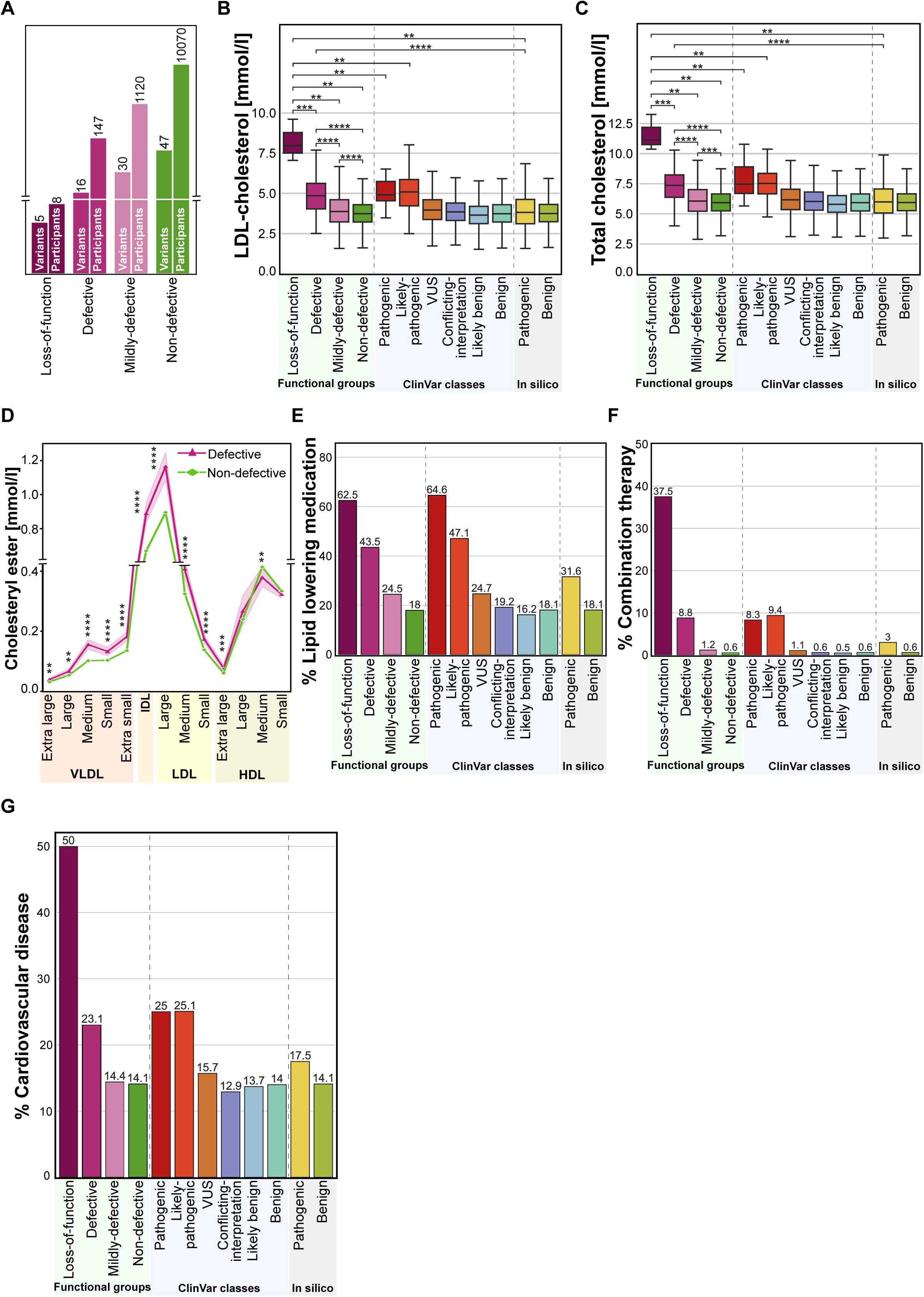
Integrating functional groups of *LDLR* variants with whole-exome sequencing and clinical data from UK Biobank. (A) Number of functionally characterized *LDLR* variants and individuals carrying these variants retrieved from UK Biobank. (B) and (C) Assessment of the impact of LDLR functional groups, along with their ClinVar and predicted classifications, on circulating LDL cholesterol and total cholesterol levels, respectively in participants who are not receiving lipid-lowering medication or combination therapy. (D) Effect of functional groups on lipoprotein composition. (E) and (F) Comparative analysis of the utilization of lipid-lowering medication and combination therapy, respectively, by individuals carrying different classes of LDLR variants. (G) The proportion of individuals experiencing cardiovascular events within each LDLR variant group categorized by distinct classification systems. Statistical significance was determined using the Mann-Whitney U test, with asterisks denoting significance levels: *p < 0.05, **p < 0.01, ***p < 0.001 and ****p < 0.0001.

**Table 1:** List of odds ratios related to Figure 6.

| Events | Variant classes | Odds ratio | 95% Confidence intervals | p-value |
| --- | --- | --- | --- | --- |
| **LDL cholesterol**<br>**≥ 5 mmol/L**<br>**(UK Biobank)** | Loss-of-function/Non-defective | 15.4 | 3.8 - 61.7 | 8.0e-04 |
|  | Defective/Non-defective | 5.9 | 4.1 - 8.6 | 2.5e-16 |
|  | Mildly defective/Non-defective | 1.6 | 1.3 - 2.0 | 1.6e-05 |
|  | ClinVar Pathogenic/Benign | 5.1 | 2.6 - 9.9 | 2.6e-05 |
|  | ClinVar Likely pathogenic/Benign | 6.4 | 4.7 - 8.9 | 5.9e-23 |
| **Lipid-lowering**<br>**treatment**<br>**(UK Biobank)** | Loss-of-function/Non-defective | 7.6 | 1.8 - 31.9 | 6.5e-03 |
|  | Defective/Non-defective | 3.5 | 2.5 - 4.9 | 1.1e-12 |
|  | Mildly defective/Non-defective | 1.5 | 1.3 - 1.7 | 2.6e-07 |
|  | ClinVar Pathogenic/Benign | 8.3 | 4.6 - 15.0 | 1.8e-12 |
|  | ClinVar Likely pathogenic/Benign | 4.9 | 3.7 - 6.5 | 1.9e-25 |
| **Combination**<br>**therapy**<br>**(UK Biobank)** | Loss-of-function/Non-defective | 96.8 | 22.6 - 414 | 1.4e-05 |
|  | Defective/Non-defective | 15.6 | 8.4 - 29.1 | 3.9e-11 |
|  | Mildly defective/Non-defective | 1.9 | 1.0 - 3.4 | 4.9e-02 |
|  | ClinVar Pathogenic/Benign | 15.9 | 5.4 - 46.0 | 2.0e-04 |
|  | ClinVar Likely pathogenic/Benign | 21.3 | 12.4 - 36.5 | 8.8e-19 |
| **Cardiovascular**<br>**risk**<br>**(UK Biobank)** | Loss-of-function/Non-defective | 6.1 | 1.5 - 24.4 | 1.7e-02 |
|  | Defective/Non-defective | 1.8 | 1.2 - 2.7 | 3.8e-03 |
|  | Mildly defective/Non-defective | 1 | 0.8 - 1.2 | 7.8e-01 |
|  | ClinVar Pathogenic/Benign | 2 | 1.0 - 3.9 | 3.7e-02 |
|  | ClinVar Likely pathogenic/Benign | 2.1 | 1.5 - 3.0 | 9.1e-06 |

However, it became apparent that especially in the loss-of-function group a mix of recipients with lipid-lowering therapy (Supplementary Fig. 2e) and naive patients (Fig. 6b) led to a large spread of LDL-C values (Supplementary Fig. 2b). As expected, the highest values for circulating LDL-C were observed in subjects without lipid-lowering treatment (Fig. 6b), with LDL-C values for the loss-of-function group (7.96 mmol/l) being more than two-fold higher than for the non-defective variant group. LDL-C values were also higher in the defective (4.99 mmol/l), and mildly-defective variant groups (3.87 mmol/l) than for the non-defective variant group (3.74 mmol/l) (Fig. 6b). Similar observations were made for plasma concentrations of total cholesterol (11.12 mmol/l for the loss-of-function, 7.45 mmol/l for the defective, 6 mmol/l for the mildly-defective and 5.92 mmol/l for the non-defective group) (Fig. 6c) and for apolipoprotein B (APO-B) (Supplementary Fig. 2h). Interestingly, LDL-C and total cholesterol values were lower for the ClinVar pathogenic and likely-pathogenic variants as compared to the loss-of-function group. Furthermore, LDL-C, total cholesterol and APO-B were higher in loss-of-function and defective groups as compared to the predicted pathogenic group and also the VUS group (Fig. 6b, c, Supplementary Fig. 2h). Functional impairment of the LDLR also affected the composition and other lipoprotein classes, as highlighted by an increased cholesterol ester content in small VLDL, IDL and several LDL size-classes for the defective *LDLR* variant group as compared to the non-defective variant group, for subjects without lipid-lowering medication (Fig. 6d).

Moreover, we observed differences in the utilization of lipid-lowering drugs for the functional activity groups. More than 50% of individuals of the loss-of-function (OR = 7.6, CI: 1.8 - 31.9, p = 0.0065) and the defective group (OR = 3.5, CI: 2.5-4.9, p < 0.0001) received lipid-lowering drugs (Fig. 6e, Table 1). A further differentiation revealed that 37.5% of the loss-of-function group (OR = 96.8, CI: 22.6 - 414, p = 0.0001) received combination lipid-lowering therapy, 11% for the defective group (OR = 15.6, CI: 8.4 - 29.1, p < 0.0001), 1.2% for the mildly-defective group (OR = 1.9, CI: 1.0-3.4, p = 0.049) and 0.6% of the non-defective group (Fig. 6f, Table 1).

Next, we investigated the effect of functional *LDLR* variant groups on cardiovascular events for the UKB subjects. 50% of carriers of a loss-of-function variant experienced a cardiovascular event, 23.1% of those with a defective variant, 14.4 % with a mildly-defective variant and 14.0% of those carrying a variant from the non-defective group (Fig. 6g). The corresponding odds ratios for experiencing a cardiovascular event as compared to subjects from the non-defective group were 6.1 (CI: 1.5 - 24.4, p = 0.017) for the loss-of-function group and 1.8 (CI 1.2 - 2.7, p = 0.0038) for the defective group (Table 1).

## Discussion

We designed a systematic and semi-automated approach to overcome current limitations in functional profiling of *LDLR* variants. At the same time, we desired a screening system which is more close to human physiology compared to previous efforts^9,10^. Therefore, we utilized a human liver cell line (HepG2) in which we disrupted LDLR activity with CRISPR and then reintroduced LDLR-GFP expression constructs into the genome at the AAVS1 safe harbor locus. Compared to heterologous systems to assess LDLR activities^8^, our approach has several advantages. 1) A cell line of liver origin increases the likelihood that a relevant set of adaptor proteins for efficient LDLR trafficking is present. 2) Subtle changes in 3D structure of adaptor proteins due to sequence differences in non-human cell systems may alter LDLR trafficking. Our system overcomes this limitation. 3) Stable genomic integration of LDLR-GFP variant expression enables quantification of a large number of cells in each experiment. 4) Genomic integration allows low-level and homogenous expression of the LDLR-GFP construct, leading to more precise quantification of altered LDLR-GFP protein expression. For example we showed increased wild-type LDLR-GFP expression in lipid poor conditions as compared to lipid rich conditions. This effect was further enhanced by inhibiting cholesterol synthesis, highlighting that additional regulatory factors can influence LDLR protein expression beyond transcriptional control of LDLR mRNA. The underlying mechanisms may involve inducible degradation of LDLR^24^ or mechanisms involving receptor recycling^25,26^, but requires more research to obtain a deeper understanding.

A defective *LDLR* variant comes along with lower LDL internalization and hence intracellular cholesterol depletion. This may lead to cellular adaptations which stabilize the LDLR protein. Our observations for increased LDLR-GFP expression for ligand-binding domain variants supports this. On average, higher LDLR-GFP protein expression was not observed for mutations from other LDLR domains. Probably, these mutations directly affect the balance of receptor sorting and degradation by changing affinities to adaptor proteins or destabilizing the protein itself.

Prediction tools can be used to estimate the pathogenicity of *LDLR* variants. Often *LDLR* variants are considered pathogenic if they are labeled defective by multiple prediction programs^4^. In our case, only a subset of predicted pathogenic *LDLR* variants represented loss-of-function or defective variants. This highlights limitations of prediction tools, which seem to overestimate the amount of pathogenic variants and thereby influence cardiovascular risk estimations and calculations of FH prevalence.

Interestingly, some missense variants showed a clear loss-of-function phenotype even though, in general, they are considered to lead to less severe consequences. Pinpointing carriers of such *LDLR* variants will be important for effective cardiovascular risk reduction. Moreover, the functional groups provide more in-depth information than the ClinVar groups as highlighted by our UK Biobank analysis regarding circulating lipoproteins, utilization of lipid-lowering drugs and cardiovascular risk. For example, the loss-of-function group resulted in more severe phenotypes as compared to the ClinVar groupings for the same variants and may therefore enable the identification of FH patients with profound cardiovascular risk and a pressing need for effective pharmaceutical intervention. Grouping *LDLR* variants into additional defective and mildly-defective groups allowed us to demonstrate differential effects of these variant groups on dyslipidaemia and CVD progression. Consequently, more precise risk assessment can be performed for a person, going beyond binary variant groups such as pathogenic or benign. Therefore, our study provides valuable data for precision medicine applications in FH. In this context, expanding our analysis to additional *LDLR* variants will be of key importance.

Currently, high-content assays can only support the classification of *LDLR* variants as they are labeled as class 3 assays (PS3_Supporting/BS3_Supporting) by the ClinGen FH VCEP expert panel^6^. Increasing the assay class of high-content experiments to class 2 or 1 would lead to a higher impact of functional cell data on clinical decision making. In our analysis platform, we included positive (KO-LDLR-WT-GFP) and negative LDLR KO controls in every experiment and quantified variants in multiple replicates in two independent experiments. According to the VCEP classification rules (pathogenic variant = 0-70% residual LDLR activity) 83% of ClinVar pathogenic variants were detected as pathogenic by our platform and none as benign, whilst 75% of the ClinVar benign variants were validated as benign. Consequently, our data provides additional evidence for increasing the classification level of high-content assays.

We observed a large number of carriers of mildly-defective *LDLR* variants in the UK Biobank. Whilst the effect on plasma lipids, utilization of lipid-lowering medication and cardiovascular events was less pronounced for these variants as compared to defective or loss-of-function variants, it is likely that mildly-defective variants influence disease progression through interaction with other gene variants from the same pathway. Therefore, precise functional information for more gene variants opens up unique opportunities to demystify the complex genetics of dyslipidaemia.

### Limitations of the study

Only a limited number of subjects with loss-of-function variants were contained in UK biobank. This might impact the magnitude of plasma lipid alterations and odds ratios for cardiovascular disease. On the other hand, our data aligns with genetic studies showing that loss-of-function *LDLR* variants result in higher cardiovascular risk^4,27,28^. The UK Biobank represents mostly white individuals with a European background. Valuable insight could be gained by assessing the effect of LDLR activity groups on dyslipidaemia and CVD in subjects from additional ethnic groups.

Our study provides an unprecedented resource for *LDLR* missense, frameshift and indel variants, assigning residual LDL uptake activities to 315 variants. This brings along benefits for early diagnosis of FH, improves risk assessment of hypercholesterolaemia and cardiovascular disease and may influence the selection of treatment strategies in FH. Functional activity scores for hypercholesterolaemia gene variants may change the way we conceive the interrelated contribution of monogenic, polygenic and so far uncharacterized gene variants to the progression of hypercholesterolaemia and cardiovascular disease.

## Data Availability

All data produced in the present study are available upon reasonable request to the authors

https://www.ukbiobank.ac.uk/

## Acknowledgements

We would like to thank Valeria Ullrich and Sara Ranta for their technical assistance in the generation of LDLR variant expression constructs and cell culture.

This research has been conducted using data from UK Biobank a major biomedical database, with the permission number: 87446 https://www.ukbiobank.ac.uk/

## Data availability statement

Imaging data will be made available on online platforms and data and code is available from the authors upon request.

---

**Supplementary Figure 1:** Verification of LDLR KO cell line. (A) Sequence analysis of the targeted amplified region of *LDLR* from LDLR KO cell lines in Exon 6 demonstrating insertion modifications in both *LDLR* alleles. (B) Cell surface expression levels of LDLR in wild-type HepG2, LDLR KO, and LDLR WT-EGFP expressing KO cell lines. (C) Overall cell expression (internal + surface) of LDLR in wild-type HepG2, LDLR KO, and LDLR WT-EGFP expressing KO cell lines. (D) Density plot of GFP intensities for individual cells of GFP-tagged LDLR variants cell lines compared to KO-LDLR-WT-GFP cell lines. Approximately 80000 cells were quantified from 5 independent experiments. Quantification of LDLR expression was performed using two independent experiments, each with duplicate wells for each treatment condition. Statistical significance was determined using the Mann-Whitney U test, with asterisks denoting significance levels: *p < 0.05, **p < 0.01, ***p < 0.001 and ****p < 0.0001.

**Supplementary Figure 2:** (A) Higher expression of variants located in the ligand-binding domain of LDLR. Comparative analysis of the levels of (B) LDL cholesterol, (C) total cholesterol, and (D) Apolipoprotein B (APO-B) in UK Biobank participants, irrespective of their lipid-lowering medication status, across different LDLR classification systems. Comparative analysis of the levels of (E) LDL cholesterol, (F) total cholesterol, and (G) APO-B in UK Biobank participants who are on lipid-lowering medication or combination therapy, across different LDLR classification systems. (H) Levels of APO-B in UK Biobank participants who are not receiving lipid-lowering therapy or combination therapy. Statistical significance was determined using the Mann-Whitney U test, with asterisks denoting significance levels: *p < 0.05, **p < 0.01, ***p < 0.001 and ****p < 0.0001.

**Supplementary Table 1:** Information on *LDLR* variants and their functional data.

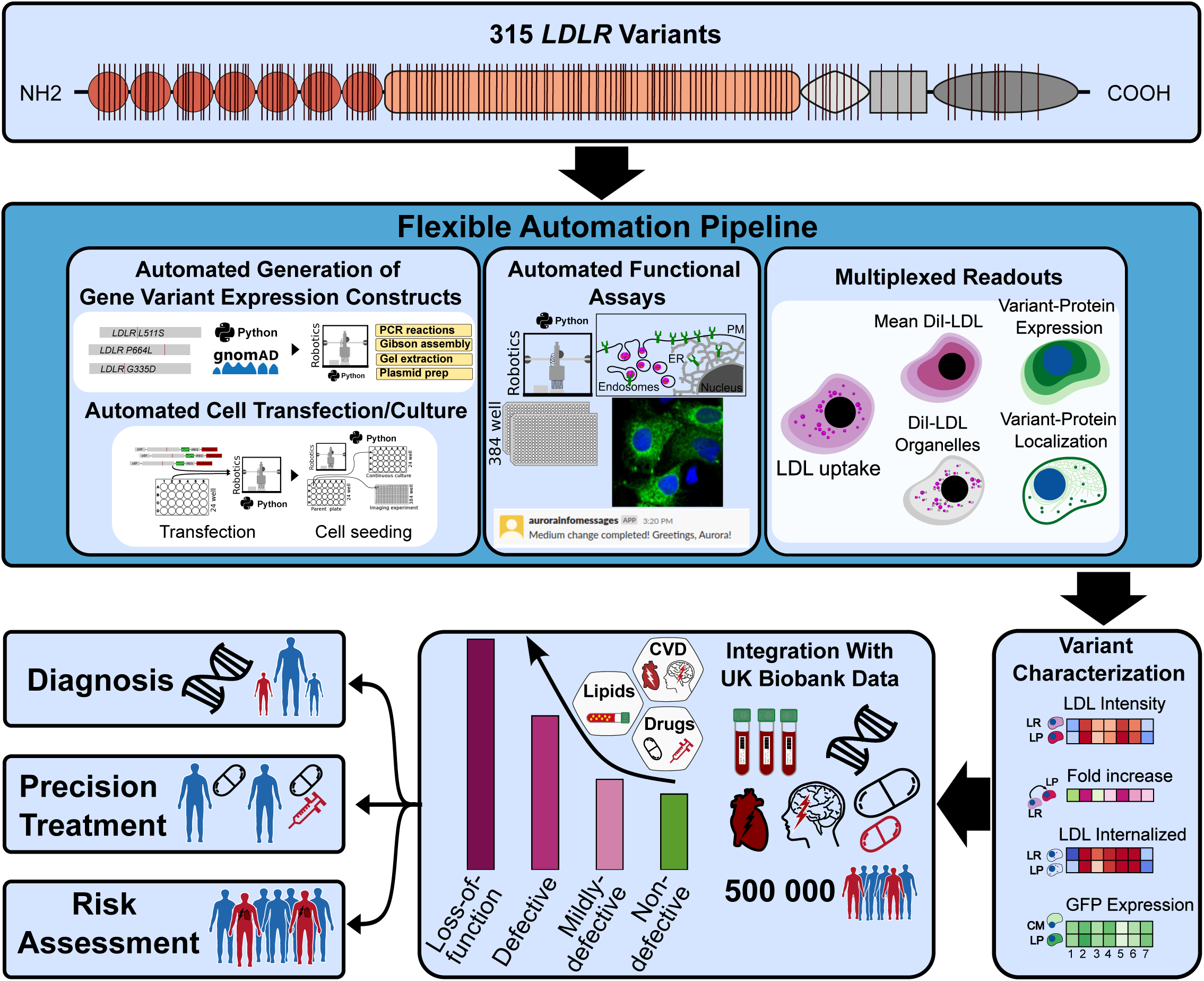

## References

1. Borén, J. et al. Low-density lipoproteins cause atherosclerotic cardiovascular disease: pathophysiological, genetic, and therapeutic insights: a consensus statement from the European Atherosclerosis Society Consensus Panel. Eur. Heart J. (2020) doi:10.1093/eurheartj/ehz962.

2. Mach, F. et al. 2019 ESC/EAS guidelines for the management of dyslipidaemias: Lipid modification to reduce cardiovascular risk. Atherosclerosis 290, 140–205 (2019).

3. Nordestgaard, B. G. et al. Familial hypercholesterolaemia is underdiagnosed and undertreated in the general population: guidance for clinicians to prevent coronary heart disease: Consensus Statement of the European Atherosclerosis Society. Eur. Heart J. 34, 3478–3490 (2013).

4. Khera, A. V. et al. Diagnostic Yield and Clinical Utility of Sequencing Familial Hypercholesterolemia Genes in Patients With Severe Hypercholesterolemia. J. Am. Coll. Cardiol. 67, 2578–2589 (2016).

5. Berberich, A. J. & Hegele, R. A. The complex molecular genetics of familial hypercholesterolaemia. Nat. Rev. Cardiol. 16, 9–20 (2019).

6. Chora, J. R. et al. The Clinical Genome Resource (ClinGen) Familial Hypercholesterolemia Variant Curation Expert Panel consensus guidelines for LDLR variant classification. Genet. Med. (2021) doi:10.1016/j.gim.2021.09.012.

7. Bedlington, N., et al. The time is now: Achieving FH paediatric screening across Europe – The Prague Declaration. GMS Health Innov. Technol. 16, Doc04 (2022).

8. Bourbon, M., Alves, A. & Sijbrands, E. Low-density lipoprotein receptor mutational analysis in diagnosis of familial hypercholesterolemia. Curr. Opin. Lipidol. 28, 120–129 (2017).

9. Thormaehlen, A. S. et al. Systematic Cell-Based Phenotyping of Missense Alleles Empowers Rare Variant Association Studies: A Case for LDLR and Myocardial Infarction. PLOS Genet. 11, e1004855 (2015).

10. Graça, R., Zimon, M., Alves, A. C., Pepperkok, R. & Bourbon, M. High-Throughput Microscopy Characterization of Rare LDLR Variants. JACC Basic Transl. Sci. 8, 1010–1021 (2023).

11. Findlay, G. M., Boyle, E. A., Hause, R. J., Klein, J. C. & Shendure, J. Saturation editing of genomic regions by multiplex homology-directed repair. Nature 513, 120–123 (2014).

12. Findlay, G. M. et al. Accurate classification of BRCA1 variants with saturation genome editing. Nature 562, 217–222 (2018).

13. Fowler, D. M. & Fields, S. Deep mutational scanning: a new style of protein science. Nat. Methods 11, 801–807 (2014).

14. Hanna, R. E., et al. Massively parallel assessment of human variants with base editor screens. Cell 184, 1064-1080.e20 (2021).

15. Lue, N. Z. & Liau, B. B. Base editor screens for in situ mutational scanning at scale. Mol. Cell 83, 2167–2187 (2023).

16. Li, S., Prasanna, X., Salo, V. T., Vattulainen, I. & Ikonen, E. An efficient auxin-inducible degron system with low basal degradation in human cells. Nat. Methods 16, 866–869 (2019).

17. Anderson, R. G., Brown, M. S., Beisiegel, U. & Goldstein, J. L. Surface distribution and recycling of the low density lipoprotein receptor as visualized with antireceptor antibodies. J. Cell Biol. 93, 523–531 (1982).

18. Gibson, D. G. et al. Enzymatic assembly of DNA molecules up to several hundred kilobases. Nat. Methods 6, 343–345 (2009).

19. Ng, P. C. & Henikoff, S. SIFT: predicting amino acid changes that affect protein function. Nucleic Acids Res. 31, 3812–3814 (2003).

20. Steinhaus, R. et al. MutationTaster 2021. Nucleic Acids Res. 49, W446–W451 (2021).

21. Adzhubei, I., Jordan, D. M. & Sunyaev, S. R. Predicting Functional Effect of Human Missense Mutations Using PolyPhen-2. Curr. Protoc. Hum. Genet. Editor. Board Jonathan Haines Al 0 7, Unit7.20 (2013).

22. Capriotti, E. & Fariselli, P. PhD-SNPg: a webserver and lightweight tool for scoring single nucleotide variants. Nucleic Acids Res. 45, W247–W252 (2017).

23. Cheng, J. et al. Accurate proteome-wide missense variant effect prediction with AlphaMissense. Science 381, eadg7492 (2023).

24. Zelcer, N., Hong, C., Boyadjian, R. & Tontonoz, P. LXR Regulates Cholesterol Uptake Through Idol-Dependent Ubiquitination of the LDL Receptor. Science 325, 100–104 (2009).

25. Wijers, M., Kuivenhoven, J. A. & van de Sluis, B. The life cycle of the low-density lipoprotein receptor: insights from cellular and in-vivo studies. Curr. Opin. Lipidol. 26, 82–87 (2015).

26. Rimbert, A. et al. A common variant in CCDC93 protects against myocardial infarction and cardiovascular mortality by regulating endosomal trafficking of low-density lipoprotein receptor. Eur. Heart J. 41, 1040–1053 (2020).

27. Trinder, M., Francis, G. A. & Brunham, L. R. Association of Monogenic vs Polygenic Hypercholesterolemia With Risk of Atherosclerotic Cardiovascular Disease. JAMA Cardiol. 5, 390–399 (2020).

28. Clarke, S. L. et al. Coronary Artery Disease Risk of Familial Hypercholesterolemia Genetic Variants Independent of Clinically Observed Longitudinal Cholesterol Exposure. Circ. Genomic Precis. Med. 15, e003501 (2022).

