## Supplementary Methods for "Large-scale functional characterization of low-density lipoprotein receptor gene variants improves risk assessment in cardiovascular disease"

**Reagents**

Hoechst 33342 (Thermo Fisher, Cat: H1399), CellMask Deep Red (Thermo Fisher, Lot: 216004), anti-mouse Alexa Fluor 568 (Cat: A11004), Lipofectamine LTX & PLUS reagents (Cat: 15338030), Dil Stain (1,1'-Dioctadecyl-3,3,3',3'-Tetramethylindocarbocyanine Perchlorate ('DiI'; DiIC18(3))) (Cat: D3911) and fetal bovine serum (FBS) (Cat: 10270106) were purchased from Thermo Fisher. Q5 Hot Start High-Fidelity DNA polymerase (Cat: M04932), Phusion High-Fidelity Polymerase (Cat: F530S), Taq DNA ligase (Cat: M0208S), T5 Exonuclease (Cat: M0363S), Bgl II (Cat: ER0081), EcoRI-HF (Cat: R3101S) were purchased from New England Biolabs. L-glutamate (Cat: BE17-605E) and Penicillin streptomycin (Cat: DE17-602E) from Lonza. Anti-hLDLR mouse monoclonal antibody (clone 472413, Cat: MAB24) was purchased from R&D systems, Poly-L-lysine (Cat: 11426642 ) from MP Biochemicals, Mevastatin (Cat: M2537-5MG) from Sigma-Aldrich, RPMI-1640 (Cat: ECB9006L) from EuoClone, SeraSil-Mag 400 silica coated superparamagnetic beads (Cat: 29357371) from Cytiva, and dNTPs (Cat: U1240) were purchased from Promega.

Lipoprotein-deficient serum (LPDS), DiI-labeled low-density lipoprotein (DiI-LDL) were produced as previously described^16,17^.

**Cell culture and generation of the LDLR knockout cell line**

HepG2 cells were purchased from the European Collection of Authenticated Cell Cultures ECACC and cultured cultured in RPMI 1640 supplemented with 10% FBS, 1% L-glutamate, and 1% Penicillin streptomycin.

LDLR knockout (LDLR KO) cells were generated using the CRISPR-Cas9 technology as described previously. Two guide RNAs (gRNAs) specific to the LDLR gene were designed using online tools (<http://chopchop.cbu.uib.no/>) and cloned into plasmids carrying the Cas9 nickase gene. These were transfected into HepG2 cells and knockout cell lines, derived from a single clone, were pre-screened with PCR-PAGE^18^ and validated with Sanger sequencing.

**Selection of LDLR variants and generation of LDLR variant expression vectors**

We selected five pathogenic, and three benign variants together with 35 ClinVar likely-pathogenic variants to establish the workflow and to validate our approach. Then we selected 400 *LDLR* variants randomly from ClinVar and the Genome aggregation database (gnomAD, https://gnomad.broadinstitute.org/). The Ensembl *LDLR* transcript ENST00000558518.6 (RefSeq NM_000527.4, UniProt ID P01130-1) which encodes a protein of 860 amino acids was utilized as the reference sequence for annotation of the *LDLR* variants.

A custom-designed Python script was used to automate the design of a large number of oligonucleotides for generating LDLR variants (https://zenodo.org/records/10359715)^19^ and were ordered from Eurofins Genomics. The oligonucleotides were used to amplify two LDLR fragments which contained the desired mutation in an overlapping region. The fragments were purified and Gibson assembled with the pSH-EFIRES-P vector backbone^20,21^, opened with Bgl2 and Not1 restriction enzymes, and transformed into DH5 alpha competent cells.

**DNA isolation**

Plasmid isolation and gel extraction of PCR products was performed on an Opentrons OT-2 robotic platform utilizing a protocol from Oberacker et al.^22^.

**Generation of variant cell lines**

GFP-tagged LDLR WT and LDLR variants were stably expressed in LDLR KO cells using CRISPR-Cas9 technology. The constructs were targeted to the adeno-associated virus integration site 1 (AAVS1) safe harbor locus of LDLR KO cells under and constitutively expressed by the EF1-alpha promoter. Plasmids harboring the LDLR WT gene and LDLR variants were co-transfected with a Cas9 plasmid into the LDLR KO cells using lipofectamine LTX and PLUS reagents. For transfection, LDLR KO cells were seeded in 24-well plates at a density of 300 000 cells per well and stable cell lines were selected with puromycin. Transfection procedures were automated and performed using an OT-1 robotic platform (Opentrons).

**Image analysis and quantification**

A custom-made Python tool (<https://github.com/lopaavol/OPutils>)^16,17,23^ was used to collapse z-stack images into maximum intensity projections. The nuclei, cytoplasm and endocytic organelles filled with DiI-LDL were automatically segmented and quantified using CellProfiler^24,25^. Cellular DiI-LDL intensity, number of DiI-LDL positive endosomal organelles and cellular GFP intensity were quantified for each cell. On average, over 100 cells were quantified from each image field within each well. This resulted in the quantification of over 1,500 cells for each cell line and each treatment condition across all assays performed.

**Prediction tools**

Five in silico tools were used to predict the effect of *LDLR* variants: PolyPhen-2^16^ (two classifier models: HumVar and HumDiv, <http://genetics.bwh.harvard.edu/pph2>), MutationTaster^17^ (<https://www.genecascade.org/MutationTaster2021/>), PhD-SNPg^18^(<https://snps.biofold.org/phd-snpg/index.html>), SIFT^19^ (<https://sift.bii.a-star.edu.sg/index.html>). Variants that received a pathogenic classification from all prediction tools were considered as predicted pathogenic. Three of our frameshift indel variants were excluded from the prediction analysis and five nonsense variants included in this study could not be analyzed by PolyPhen-2.
