## Supplementary figures and images for "Large-scale functional characterization of low-density lipoprotein receptor gene variants improves risk assessment in cardiovascular disease"

### Supplementary Figure 1

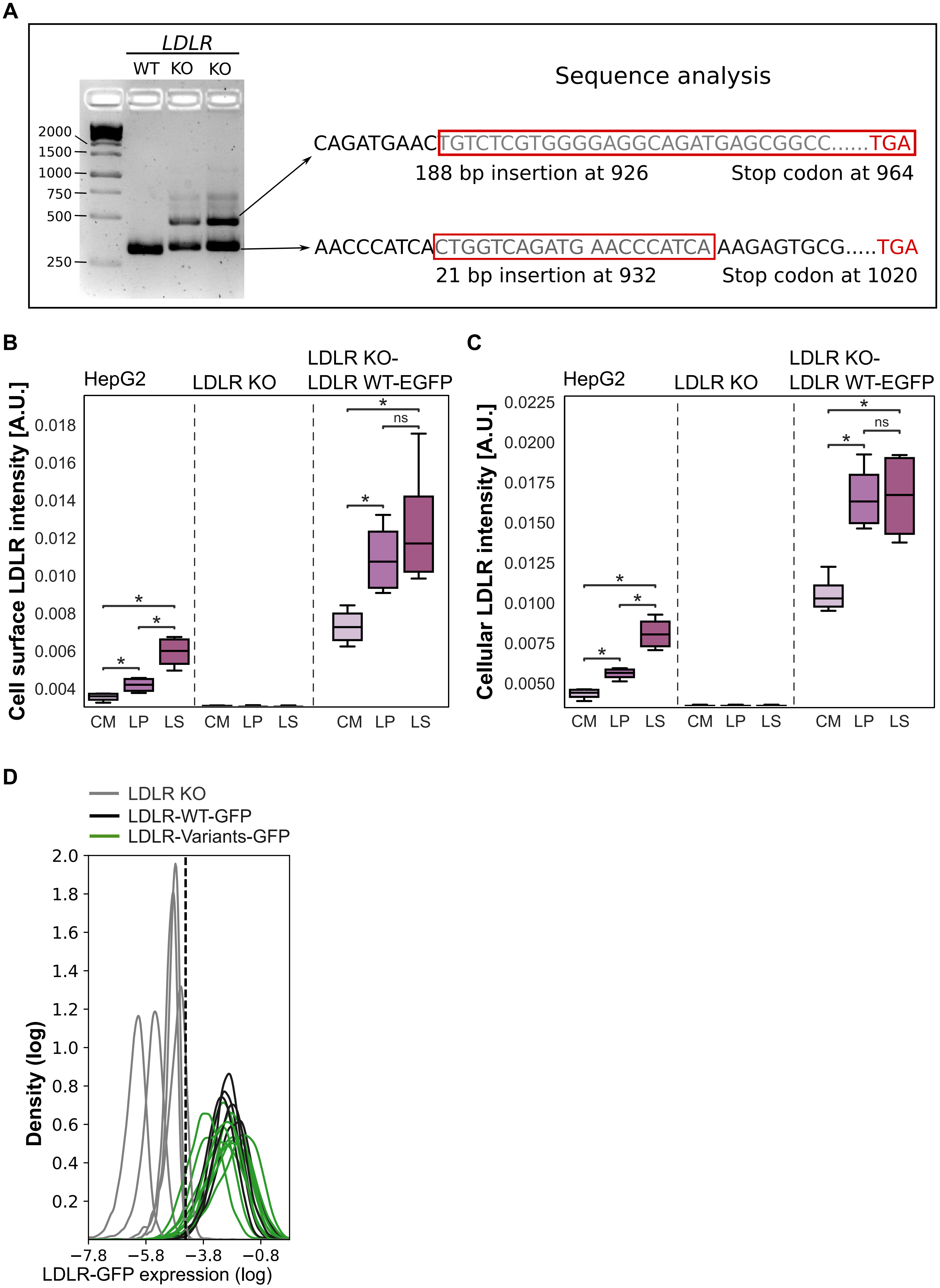


**Supplementary Figure 1**

### Supplementary Figure 2

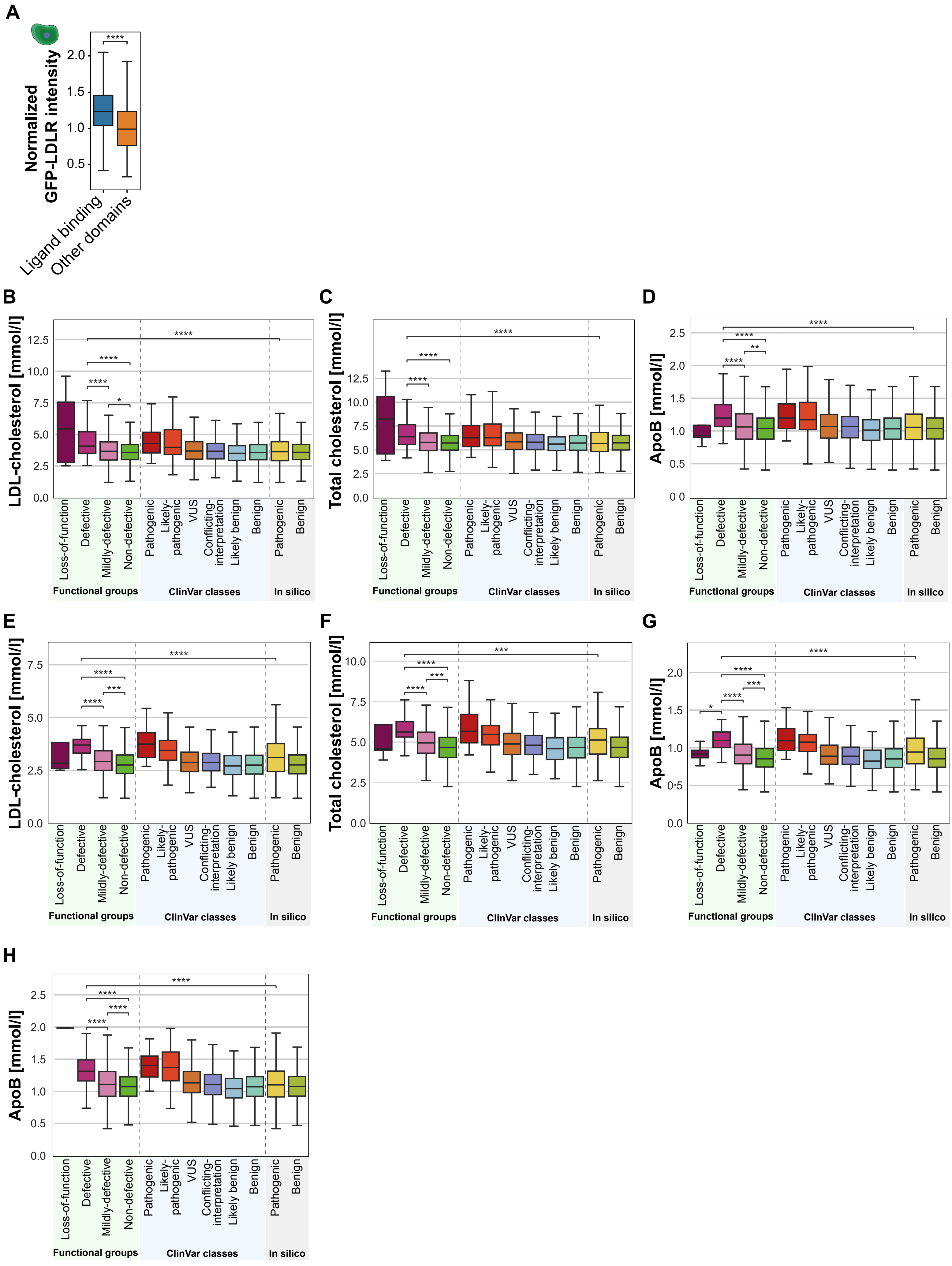


**Supplementary Figure 2**
